# CRISPR-Cas12a Assay for Rapid and Specific Detection of *Shigella flexneri 2a* in Clinical Samples

**DOI:** 10.64898/2025.12.19.25342683

**Authors:** Jipei Liao, Yun Su, Feng Jiang

## Abstract

*Shigella flexneri* 2a is the most common cause of shigellosis, a major public health concern in developing countries. Rapid and reliable diagnostic tools are critical for timely outbreak detection and management. Leveraging Clustered Regularly Interspaced Short Palindromic Repeats (CRISPR) technology, we developed a CRISPR-Cas12a-based assay for the rapid and specific detection of *S. flexneri* 2a and validated its performance using stool specimens from patients. Two guide RNAs targeting the *gtrII* and *gtrX* genes-unique markers of the *S. flexneri* 2a serotype-were designed to ensure specificity. Recombinase Polymerase Amplification (RPA) was coupled with Cas12a-mediated collateral cleavage for signal amplification, with detection by fluorescence or lateral flow. Analytical sensitivity, specificity, and clinical accuracy were compared with conventional PCR using purified DNA and 588 clinical stool specimens. The CRISPR-Cas12a assay achieved a detection limit of 10 copies/uL, comparable to PCR, and showed 100% analytical specificity without cross-reactivity to other bacteria. The isothermal reaction operated at room temperature and was completed within one hour. Both readouts allowed visual interpretation without specialized equipment. Clinical validation demonstrated a diagnostic sensitivity of 95% and specificity of 98%, confirming robust performance. This study provides two key advances: it establishes a CRISPR-Cas12a assay specifically targeting *S. flexneri* 2a, the predominant serotype, and validates it using a large clinical cohort. The assay’s simplicity, speed, and high diagnostic accuracy make it a valuable tool for clinical diagnostics and field-based surveillance in resource-limited settings.

**IMPORTANCE:** Rapid and accessible diagnostics are essential for effective management of infectious diseases such as shigellosis. We developed a CRISPR-Cas12a-based assay that specifically detects *Shigella flexneri* 2a, the predominant serotype responsible for the global disease burden. This assay integrates isothermal amplification with CRISPR-mediated detection to achieve single-copy sensitivity within one hour, eliminating the need for complex instrumentation. Dual fluorescence and lateral-flow readouts enable flexible use in both clinical laboratories and low-resource settings. The method’s simplicity, accuracy, and adaptability demonstrate the practical potential of CRISPR diagnostics for point-of-care applications. By enabling rapid, on-site identification of *S. flexneri* 2a, this approach can significantly improve clinical diagnosis and strengthen public health responses to enteric pathogen outbreaks.

---

Shigellosis, an acute bloody diarrhea caused by the Gram-negative, entero-invasive bacterium *Shigella spp*., poses a significant public health challenge, particularly in developing countries. Each year, Shigella infections result in approximately 60,000 deaths among children in these regions, contributing to both acute and chronic health issues (1). The burden of *Shigella* is compounded by the rise in antimicrobial-resistant strains, which poses a challenge to current treatment strategies(1). The most prevalent strain is *S. flexneri*, particularly serotype 2a (*S. flexneri 2a*), which accounts for approximately 30-40% of *S. flexneri* infections and about one-third of all shigellosis cases in low- and middle-income countries (2). The predominance of this strain necessitates the development of effective detection methods and vaccines(3).

Current methods for detecting *S. flexneri 2a* include culture-based, serological, and PCR techniques(4). Culture-based methods, though considered the gold standard, often require several days to yield results and necessitate well-equipped laboratories and skilled personnel, which are not always available in low-resource areas (4). Serological methods, which detect antibodies or antigens related to *S. flexneri 2a*, can provide faster results but often suffer from potential false positives or false negatives. PCR-based methods offer high sensitivity and specificity and can deliver results more quickly than culture-based methods; however, they require sophisticated equipment, stable electricity, and trained laboratory technicians, limiting their widespread use in developing countries. Therefore, it is crucial to develop faster and reliable tests for *S. flexneri 2a*, which can be effective in resource-limited settings to enable timely intervention and reduce disease spread.

Clustered regularly interspaced short palindromic repeats (CRISPRs) are segments of prokaryotic DNA that feature repetitive sequences(5–7). These sequences, along with CRISPR-associated proteins (Cas), form an immune system capable of targeting and cleaving specific nucleic acids, a mechanism now utilized in gene editing(5–7). Previous studies reveal that Cas proteins can also degrade DNA and RNA, allowing ultra-sensitive nucleic acid detection(5–7). Techniques such as DETECTR and SHERLOCK, developed using Cas12a and Cas13a, have shown promise in detecting viruses like HPV, ZIKV, and DENV(5, 6). Our research demonstrates that CRISPR-Cas12a can detect a broad spectrum of viruses and genomic mutations with sensitivity comparable to that of PCR(8–12). Additionally, we developed a microplate-based CRISPR assay for simultaneous detection of multiple targets(8–13). Moreover, CRISPR has shown potential in detecting bacterial pathogens (14, 15). Cas12a-based assays can detect *Shigella flexneri* in food or artificially spiked samples; however, these methods target *S. flexneri* at the species level rather than specifically detecting *S. flexneri* 2a. Furthermore, their validation using human clinical specimens remains limited (16) (17) (18). In this study, we developed and evaluated a CRISPR-Cas12a-based assay for the rapid and specific detection of S. flexneri 2a. Compared with previous studies, our work provides two major advances: it establishes a CRISPR-Cas12a assay specifically targeting *S. flexneri* 2a, the predominant serotype, and validates its performance using human stool specimens, thereby demonstrating its clinical applicability in both healthcare and resource-limited settings.

## MATERIALS AND METHODS

### Bacterial Strains

Nucleic acid preparations of bacterial strains, including S. *flexneri, S. boydii, S. sonnei*, and *S. dysenteriae*, as well as the non-Shigella strain *E. coli* (Table 1), were obtained from the American Type Culture Collection (ATCC; Bethesda, MD). Genomic DNA isolated from the human melanoma cell line MDA-MB-435 (ATCC) was used as a negative control. The bacterial DNA was quantified and subjected to a 10-fold serial dilution in the negative control DNA, ranging from 10,000 copies/μL to 0.1 copies/μL.

**Table 1.** Bacterial Strains.

| **Table 1. Bacterial Strains** |  |  |
| --- | --- | --- |
| Organism | ATCC Number | Serotype |
| *S. boydii* | 9207 | 1 |
| *S. sonnei* | 25931 | - |
| *S. dysenteriae* | 13313 | - |
| *S. flexneri* | 700930 | 2a |
| *S. flexneri* | 12022 | 2b |
| *E. coli* | NCTC 9001 | - |

### Recombinase Polymerase Amplification (RPA) and Lateral Flow Readout of CRISPR-Cas12a Activity for Detection of S. flexneri

We performed CRISPR experiments to detect bacteria by modifying our protocols as described in previous studies [7-12]. As shown in Table 1, there are numerous serotypes of *S. flexneri*, which share highly similar or identical genomic regions. In the serotypes, the *gtrII* gene exists in both *S. flexneri 2a* and *S. flexneri 2b*, while the *gtrX* gene exists in *S. flexneri 2b, 1d, 3a, 5b*, and *X*, but not in *S. flexneri 2a*. To develop a CRISPR assay for the specific detection of *S. flexneri 2a*, we designed primers for RPA to specifically amplify the target DNA segments of *gtrII* and *gtrX*, respectively (Table 2). We then developed crRNAs that can specifically identify the sequences within the two genes (Table 2) (Figure 1).

**Table 2.** Genes Commonly Used for Identification of Various *Shigella flexneri Serotypes*.

|  |  |  |
| --- | --- | --- |
| Genes | Accession no. | Reactive *S. flexneri* serotype(s) |
| ***gtrII*** | **AY900451** | **2a, 2b** |
| ***gtrX*** | **L05001** | **1d, 2b, 3a, 5b, X** |
| *gtrV* | CP000266 | 5a, 5b |
| *wzx6* | EU294165 | 6 |
| *oac* | X59553 | 1b, 3a, 3b, 4b, 5a, 5b, 7b |
| *gtrI* | AF139596 | 1a, 1b, 1d, 7a, 7b |
| *gtrIc* | FJ905303 | 7a, 7b |
| *gtrIV* | AF288197 | 4a, 4b |
| *ipaH* | M76445 | All *Shigella* species and serotypes |
| ***Italicized bold text* shows the genes to be targeted in the CRISPR analysis for detection of *S. flexneri* 2a and 2b.** |  |  |

**Fig. 1.**
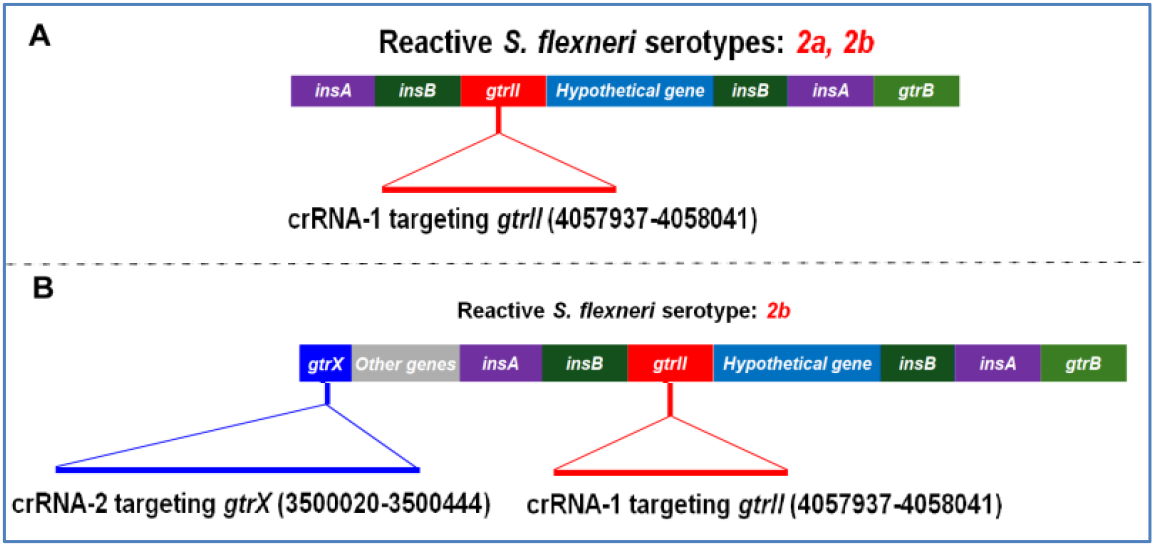
Gene Alignment and CRISPR Target Sites in *Shigella flexneri* Serotypes. A. The gene alignment in *S. flexneri 2a and 2b* and the location of crRNA-1 to specifically target the *gtrII* gene. The genes are labeled as follows: *insA, insB, gtrII, Hypothetical gene*, and *gtrB*. The crRNA-1 target region within the *gtrII* gene (base pairs 4057937-4058041) is highlighted, indicating the specific target site for crRNA-1 in the CRISPR assay. B. The gene alignment in *S. flexneri 2b* and the locations of crRNA-1 and crRNA-2 to specifically target the *gtrII* and *gtrX* genes, respectively. The genes are labeled as follows: *top, gtrB, gtrA, gtrX, gtrII*, a*rcA* and *int*. The crRNA-2 target region within the gtrX gene (base pairs 3500020-3500444) and the crRNA-1 target region within the *gtrII* gene (base pairs 4057937-4058041) are highlighted, indicating the specific target sites for crRNA-1 and crRNA-2 in the CRISPR assay.

Two crRNAs, crRNA-1 and crRNA-2 were designed for identifying and distinguishing the genes *gtrII* and *gtrX*, respectively (Table 2, Figure 1). RPA was performed by mixing 5 µL of DNA with 0.48 µM forward and reverse primers, 29.5 µL primer-free rehydration buffer, 5 µL DNA from boiled liquid samples, and 14 mM magnesium acetate (Sigma-Aldrich. St. Louis, MO) in a 50-µL reaction using the TwistAmp Basic kit (TwistDx, Cambridge, UK). The RPA mixture was incubated at 37°C for 10-20 minutes. Third, for CRSIPR reaction, 5 µL of the RPA reaction was incubated with 100 nm LbCas12a (Integrated DNA Technologies (IDT), Coralville, IA) and 100 nm crRNAs at 37°C, then diluted in binding buffer along with a dual-labeled ssDNA substrate tagged with FAM and biotin (IDT). The reaction mixture was combined with 80 µL of HybriDetect buffer (TwistDx), and a Milena HybriDetect1 lateral flow dipstick (TwistDx) was inserted into this mixture. Results were observed with the naked eye and captured using a smartphone camera. A positive result was indicated by a significant signal increase on the test line (upper line) and a decrease on the control line (lower line).

### Detection of S. flexneri 2a by CRISPR-Cas12a Using a Fluorescence Plate Reader and a UV Light Illuminator

Following RPA amplification, CRISPR-Cas12a detection was performed as described above using an ssDNA fluorophore-quencher (FQ) reporter (IDT) instead of the FAM/biotin-labeled substrate. Identical reaction mixtures were analyzed by two methods: (1) quantitative fluorescence measurement using a BioTek® Synergy™ H1 plate reader and (2) qualitative fluorescence visualization under UV illumination using a Bio-Rad ChemiDoc™ MP Imaging System (Bio-Rad Laboratories, Hercules, CA). For visual detection, fluorescence was excited under UV light, and signal intensity corresponding to target DNA presence was assessed by eye or captured photographically.

### Quantitative PCR (qPCR) Detection and Data Analysis of S. flexneri 2a

YBR Green PCR assays were performed according to previous study with modifications (19). Briefly, SYBR Green-based qPCR (50 µL) was performed using 1× PCR Master Mix, 5 µL of DNA, and gtrII/ipaH primers on a Bio-Rad CFX96 system. Cycling conditions were 95 °C for 10 min; 40-45 cycles of 95 °C for 10 s, 54.7 °C for 10 s, and 60 °C for 60 s; followed by 72 °Cfor 10 min. Melt-curve analysis confirmed specificity, and Ct ≤ 35 was considered positive. To specifically identify S. flexneri 2a, a two-gene analytical strategy was applied following the method of Liu et al (19). The ipaH gene served as a genus-level marker for all Shigella species, while the gtrII gene identified isolates belonging to serogroup 2 (*S. flexneri 2a or 2b*). A second PCR targeting the gtrX gene was used to differentiate between the two serotypes: samples positive for gtrII but negative for gtrX were classified as *S. flexneri 2a*, whereas samples positive for both gtrII and gtrX were classified as *S. flexneri 2b*. The primer sequences and diagnostic purposes are listed in Supplementary Table 1.

### Study Population and Specimens

A total of 588 stool specimens (385 *S. flexneri* 2a-positive and 203 *Shigella*-negative) were analyzed. Positive specimens were obtained from patients presenting with acute diarrhea (< 96 h duration, with mucus and ≥ 10 WBCs per high-power field) at Jiangsu Province Hospital of Chinese Medicine. All procedures were conducted under institutional review board-approved protocols, following the ethical standards. The demographic and clinical characteristics of the study population are summarized in Supplementary Table 2. The mean ± SD age of patients was 3.5 ± 1.0 years, while that of controls was 3.6 ± 0.9 years. The male-to-female ratio was approximately 1:1 in each group. All participants had no history of antibiotic exposure prior to sample collection. Infection with *Shigella flexneri* 2a was confirmed using standard bacteriological methods, including culture on MacConkey and XLD agar followed by serological identification (serotyping), as previously described (19, 20). Negative specimens included culture-negative and *ipaH/gtrII*-negative stool samples from age- and sex-matched diarrheal patients or healthy volunteers.

### Validation of CRISPR-Cas12a Assay for Detection of S. flexneri 2a in Stool Specimens

To process stool samples for nucleic acid extraction, approximately 200 mg of stool was homogenized in 1 mL of phosphate-buffered saline (Thermo Fisher, Waltham, MA) and centrifuged at 12,000 × g for 2 minutes. The supernatant was heated at 95 °C for 5 min to inactivate pathogens(21). DNA was extracted using the QIAamp Fast DNA Stool Mini Kit (Qiagen, Germany) according to previous report(22), and eluted in 200 µL AE buffer and stored at −20 °C until analysis. The CRISPR-Cas12a detection and visualization were subsequently performed as described in the previous section. Each reaction set included a positive control (*S. flexneri* 2a ATCC 700930 DNA), negative control (*E. coli* NCTC 9001 DNA), and no-template control. For comparison, all samples were also tested by qPCR as described above.

### Statistical analysis

The analytical performance of the CRISPR-Cas12a assay was evaluated using serially diluted *S. flexneri 2a* DNA (10^4^-0.1 copies/μL) to determine the limit of detection and by testing multiple Shigella species and E. coli strains to assess analytical specificity. Fluorescence intensity values were expressed as mean ± standard deviation (SD), and differences were analyzed using two-tailed Student’s t-tests, with p < 0.05 considered statistically significant. For clinical validation, based on the anticipated assay performance (sensitivity ≤ 0.90 and specificity ≤ 0.95) and a desired precision of ±3%, approximately 385 positive and 203 negative specimens were estimated to be required to achieve >90% statistical power for diagnostic validation. Sensitivity and specificity were determined from 2×2 contingency tables, and agreement with qPCR assay was assessed using Cohen’s κ coefficient. All analyses were performed using GraphPad Prism (version 10.0).

## RESULTS

### CRISPR-Cas12a Strategy for Specific Detection of S. flexneri 2a

One major challenge in the molecular detection of *Shigella* is cross-reactivity caused by sequence homology in conserved regions shared with other species and among multiple *Shigella* serotypes. In *S. flexneri*, the *gtrII* gene is present in both serotypes 2a and 2b, whereas the *gtrX* gene is found in serotypes 2b, 1d, 3a, 5b, and X, but absent in 2a. Accordingly, we designed crRNA-1 to target *gtrII* for detecting both *serotypes 2a* and *2b*, while crRNA-2 targets *gtrX* for the specific detection of *serotype 2b* (Table 3, Fig. 1). Thus, our detection strategy for *S. flexneri* 2a involves two steps (Figure. 2): first, specimens are tested with crRNA-1; positive results indicate the presence of *S. flexneri* 2a or 2b. These positives are then retested with crRNA-2-positivity confirms 2b, whereas a negative result confirms *S. flexneri* 2a

**Table 3.**
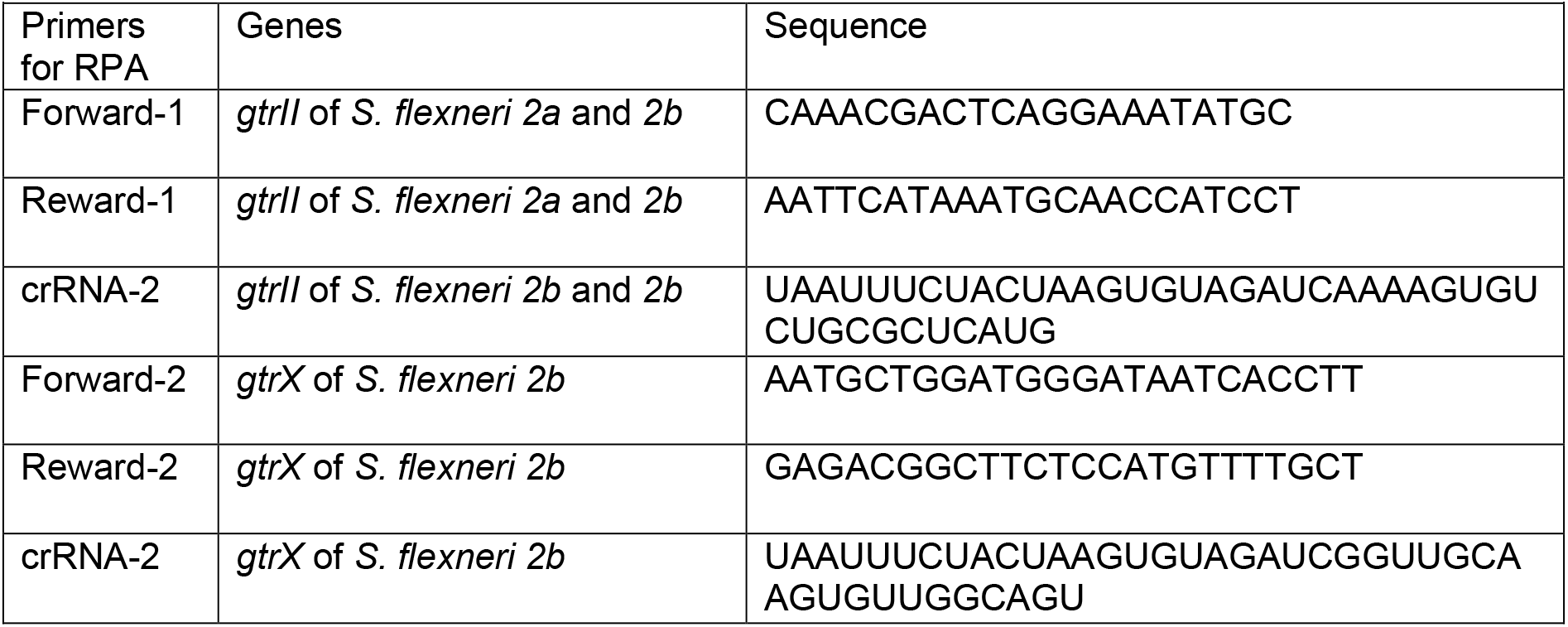
Primers for RPA and crRNAs for *Shigella flexneri* Detection.

**Fig. 2.**
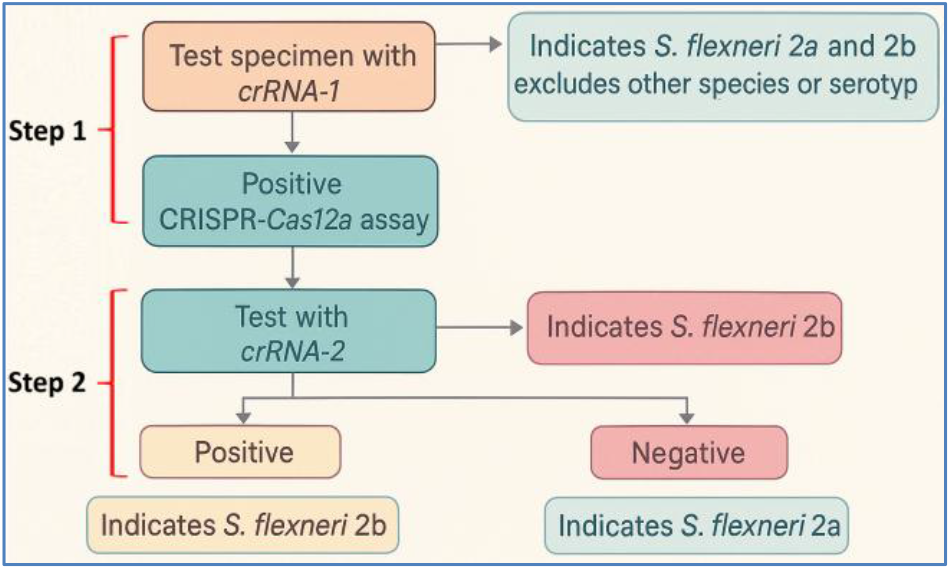
Workflow illustrating a two-step CRISPR-Cas12a assay for specific detection of *S. flexneri* 2a. Step 1: crRNA-1 targets the *gtrII* gene; a positive signal indicates the presence of *S. flexneri* 2a or 2b. Step 2: crRNA-2 targets the *gtrX* gene; a positive result confirms *S. flexneri* 2b, whereas a negative result confirms *S. flexneri* 2a.

### The CRISPR-Cas12a System with crRNAs Can Specifically Detect S. flexneri 2a

To determine the specificity of the crRNAs, CRISPR-Cas12a with the two crRNAs was tested in DNA samples from different bacterial species and serotypes (Table 1). As shown in Figure 3, CRISPR-Cas12a with crRNA-1 showed negative results in the samples of *S. boydii, S. sonnei, S. dysenteriae*, and *E. coli*. However, it showed positive results for both *S. flexneri 2a* and *2b*. The results imply that crRNA-1 is species-specific to *S. flexneri* but not serotype-specific. Interestingly, besides displaying negative results in *S. boydii, S. sonnei, S. dysenteriae*, and *E. coli*, crRNA-2 only showed a positive result in *S. flexneri 2b*, suggesting no cross-reaction of crRNA-2 between *S. flexneri 2a* and *S. flexneri 2b* (Figure 4).

**Fig. 3.**
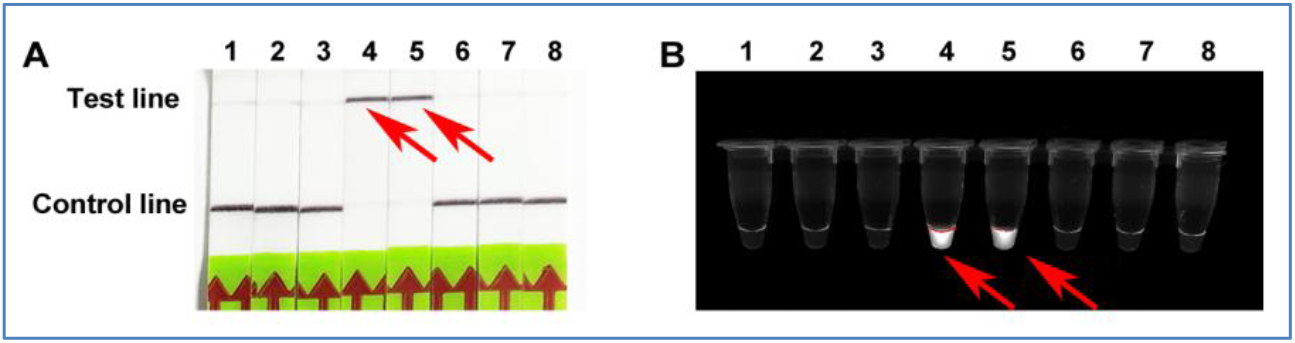
Specificity of the CRISPR assay with crRNA-1 for the detection of *S. flexneri*. A. The paper strips display the results for various bacterial strains. A positive result is indicated by a significant signal increase on the test line (upper line) and a decrease on the control line (lower line). There was a cross-reaction between *S. flexneri 2a* and *S. flexneri 2b* (Red arrows). No positive results of crRNA-1 were observed for *S. boydii 1, S. sonnei, S. dysenteriae*, and *E. coli*. Lanes: 1: *S. boydii 1*, 2: *S. sonnei*, 3: *S. dysenteriae*, 4: *S. flexneri 2a*, 5: S. *flexneri 2b*, 6: *E. coli*, 7: DNA of human MDA-MB-435 cells, and 8: NTC (no template control). B. The results of crRNA-1 for various bacterial strains were read using a UV light illuminator. Consistent with A, a positive result is indicated by a visible fluorescence signal (Red arrows). A cross-reaction between *S. flexneri 2a* and *S. flexneri 2b* was found. No positive results of crRNA-1 were observed for *S. boydii 1, S. sonnei, S. dysenteriae*, and *E. coli*.

**Fig. 4.**
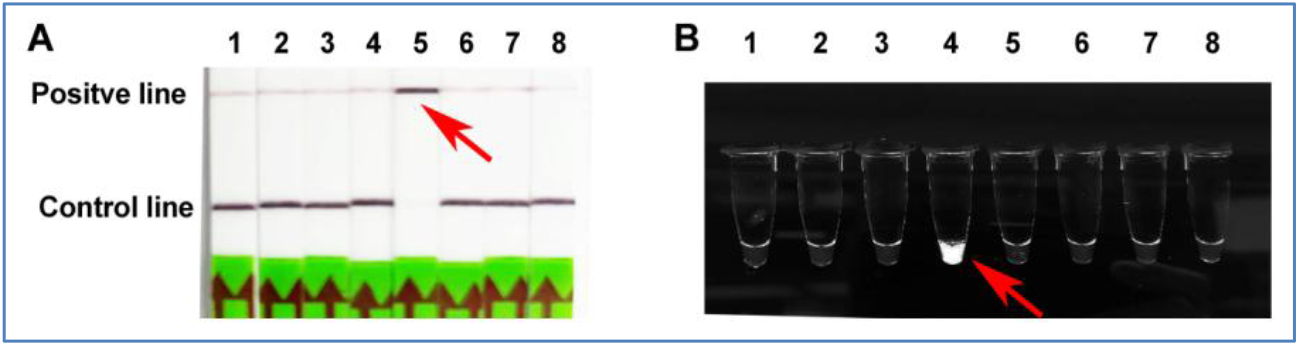
Specificity of the CRISPR assay with crRNA-2 for the detection of *S. flexneri*. A. The paper strips display that only *S. flexneri 2b* is positive for crRNA-2 (Red arrow), whereas *S. flexneri 2a* is negative for crRNA-2. No positive results were observed for other bacteria. Lanes: 1: *S. boydii 1*, 2: *S. sonnei*, 3: *S. dysenteriae*, 4: *S. flexneri 2a*, 5: S. *flexneri 2b*, 6: *E. coli*, 7: DNA of human MDA-MB-435 cells, and 8: NTC. B. The results of crRNA-2 for various bacterial strains displayed using a UV light illuminator. Only *S. flexneri 2b* (lane 4) shows a positive fluorescence signal (Red arrow), while *S. flexneri 2a* (lane 3) is negative for crRNA-2.

### The CRISPR-Cas12a System Can Sensitively Detect S. flexneri 2a

To assess the sensitivity of CRISPR-Cas12a with two crRNAs for detecting *S. flexneri 2a*, bacterial DNA was serially diluted 10-fold in DNA of human cancer cells and subjected to the CRISPR-Cas12a reaction. For comparison, the same diluted samples were also tested using PCR. Both CRISPR-Cas12a and PCR had a limit of detection of 10 copy/µL (Figure 5) (Supplementary figure 1). This parallel comparison indicates that CRISPR-Cas12a has comparable sensitivity to conventional qPCR for detecting *S. flexneri*. However, the CRISPR-Cas12a assay can be performed at room temperature within one hour, whereas PCR requires a thermocycler, different temperature cycles, and a longer reaction time of approximately three hours.

**Fig. 5.**
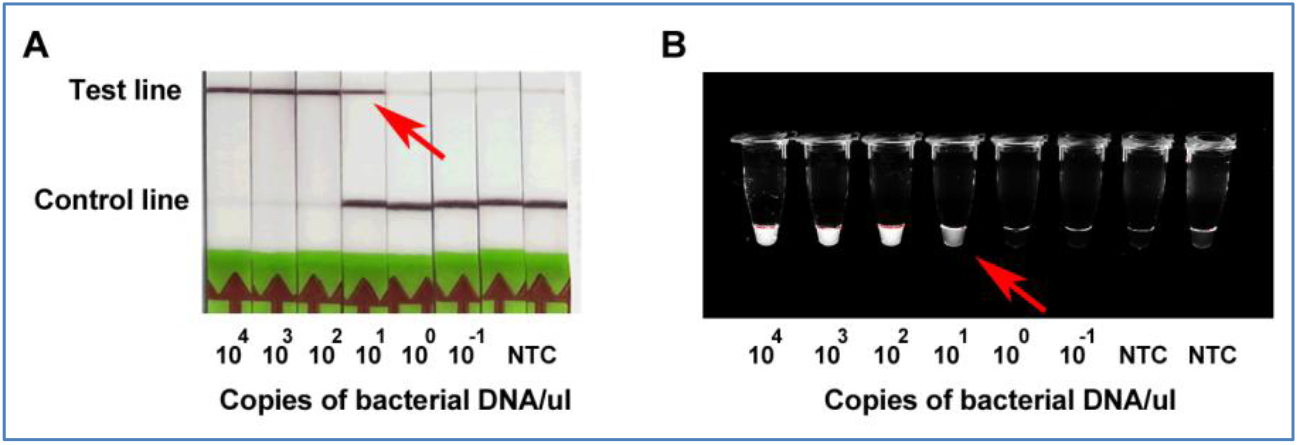
Sensitivity of the CRISPR assay for the detection of *S. flexneri*. A. The test strips display the results for various concentrations of *S. flexneri* DNA from 10,000 copies/µL to 0.1 copies/µL. The assay demonstrates the ability to detect as low as 10 copies/µL of *S. flexneri* DNA (Red arrow). B. The test tubes display the results for various concentrations of *S. flexneri* DNA, ranging from 10,000 copies/µL to 0.1 copies/µL. The assay demonstrates the ability to detect as low as 10 copies/µL of *S. flexneri* DNA (Red arrow).

### Diagnostic Performance of the CRISPR-Cas12a Assay in Clinical Specimens

Among the 385 culture-confirmed *Shigella flexneri* 2a-positive specimens, 365 were correctly detected by the CRISPR-Cas12a assay, whereas 20 were false negatives, yielding a sensitivity of 95% (Table 4). Of the 203 culture-negative control specimens, 199 were accurately classified as negative, with 4 false positives, resulting in a specificity of 98%. Furthermore, qPCR analysis demonstrates sensitivity and specificity comparable to those of the CRISPR-Cas12a assay in the same specimen set. These results confirm strong concordance between the qPCR and CRISPR-Cas12a assays and further validate the accuracy of the CRISPR-based detection platform. In addition, both fluorescence- and lateral flow-based readouts of the CRISPR-Cas12a assay yielded distinct and easily interpretable signals within 60 minutes. Moreover, the CRISPR assay requires no instrumentation, operates at room temperature, needs minimal expertise, and allows immediate visual interpretation of results with the naked eye.

**Table 4.** Diagnostic performance of the CRISPR-Cas12a assay for detection of *S. flexneri* 2a in stool specimens.

| Parameter | *S. flexneri* 2a Positive (n = 385) | *S. flexneri* 2a Negative (n = 203) | Diagnostic Measure, (95% CI) |
| --- | --- | --- | --- |
| CRISPR Positive | 365(TP) | 4(FP) | Sensitivity = 94.8%, (92.1-96.8) |
| CRISPR Negative | 20 (FN) | 199 (TN) | Specificity = 98.0%, (95.03-99.5) |

**Abbreviations:** TP = True Positive; FP = False Positive; TN = True Negative; FN = False Negative; CI = Confidence Interval.

## DISCUSSION

In this study, we demonstrate that the CRISPR-Cas12a with a pair of crRNAs can sensitively and specifically detect *S. flexneri 2a*. Furthermore, using a lateral flow dipstick or a UV light illuminator, we can rapidly visualize the results of CRISPR-Cas12a without requiring specialized technical expertise or ancillary equipment. In addition, the CRISPR-Cas12a assay showed high sensitivity and specificity comparable to qPCR (19, 23). Moreover, the strong fluorescence and lateral-flow signals obtained within 60 minutes highlight the assay’s potential as a rapid, reliable, and equipment-independent diagnostic tool for detecting *S. flexneri 2a* directly from stool specimens.

Previous reports have demonstrated the feasibility of using CRISPR-Cas12a-based platforms for the detection of *S. flexneri* in food or artificially spiked samples(16) (17) (18). However, the platforms are designed to detect *S. flexneri* in general, rather than specifically targeting *S. flexneri* 2a, which represents a major serotype, accounting for up to 40% of *S. flexneri* infections (2, 18). Furthermore, validation of CRISPR-Cas12a assays using actual human clinical specimens, particularly stool sample, has remained limited. In the present study, we developed a CRISPR-Cas12a-based detection system specifically designed to identify *S. flexneri* 2a and validated its performance using clinical stool specimens from a large cohort of cases and controls. In addition, Liu et al. developed a qPCR assay for accurate identification of *S. flexneri 2a* in clinical specimens. However, despite its high analytical performance, qPCR requires expensive instrumentation, trained personnel, and time-consuming amplification and analysis steps, limiting its use in low-resource or point-of-care settings. In contrast, our CRISPR-Cas12a assay demonstrated diagnostic accuracy comparable to qPCR in clinical samples while operating at room temperature and providing results within approximately one hour. Moreover, it enables visual detection without specialized equipment, offering a rapid, low-cost, and field-deployable alternative for effective disease surveillance and outbreak control.

Our designed crRNA-1 for identifying *S. flexneri 2a* also shows positivity for *S. flexneri 2b*. This cross-reactivity can be explained by sequence similarity, as *S. flexneri 2a* and *2b* share highly similar or identical genomic regions. However, using two distinct crRNAs in a two-step manner (Figure 2) can efficiently distinguish between *S. flexneri 2a* and *2b*, thus providing a potential diagnostic test. To further streamline detection, we plan to apply advanced bioinformatics tools to design novel crRNAs that precisely target S. flexneri 2a in a simpler and more straightforward manner.

In conclusions, this CRISPR-Cas12a assay provides a rapid, sensitive, and equipment-independent platform for the specific detection of *Shigella flexneri* 2a. Unlike previous studies that demonstrated feasibility only in food or artificially spiked samples, this work validates the performance in actual patient-derived specimens, confirming its strong potential for point-of-care applications in the prevention and management of *Shigella* outbreaks, particularly in resource-limited clinical settings.

## Data Availability

All data produced in the present work are contained in the manuscript

## Acknowledgements

We thank Dr. Søren Bentzen, Director of the Division of Biostatistics and Bioinformatics and Director of the Biostatistics Shared Service at the Greenebaum Cancer Center, for his valuable assistance with the statistical analysis.

## Author Contributions

J.L., Y.S., and F.J. conducted experiments, designed research approaches, participated in data interpretation, and prepared manuscript. All authors read and approved of the final manuscript.

## Data Availability Statement

The data that support the findings of this study are available from the corresponding author upon reasonable request.

## Institutional Review Board Statement

The study was conducted in accordance with the Declaration of Helsinki and approved by the Institutional Review Board of Nanjing University of Chinese Medicine (IRB protocol number, 23863902, approval date, April 16, 2023).

## Informed Consent Statement

Informed consent was obtained from all subjects involved in the study prior to sample

## SUPPLEMENTAL MATERIAL

**Supplementary Table 1.**
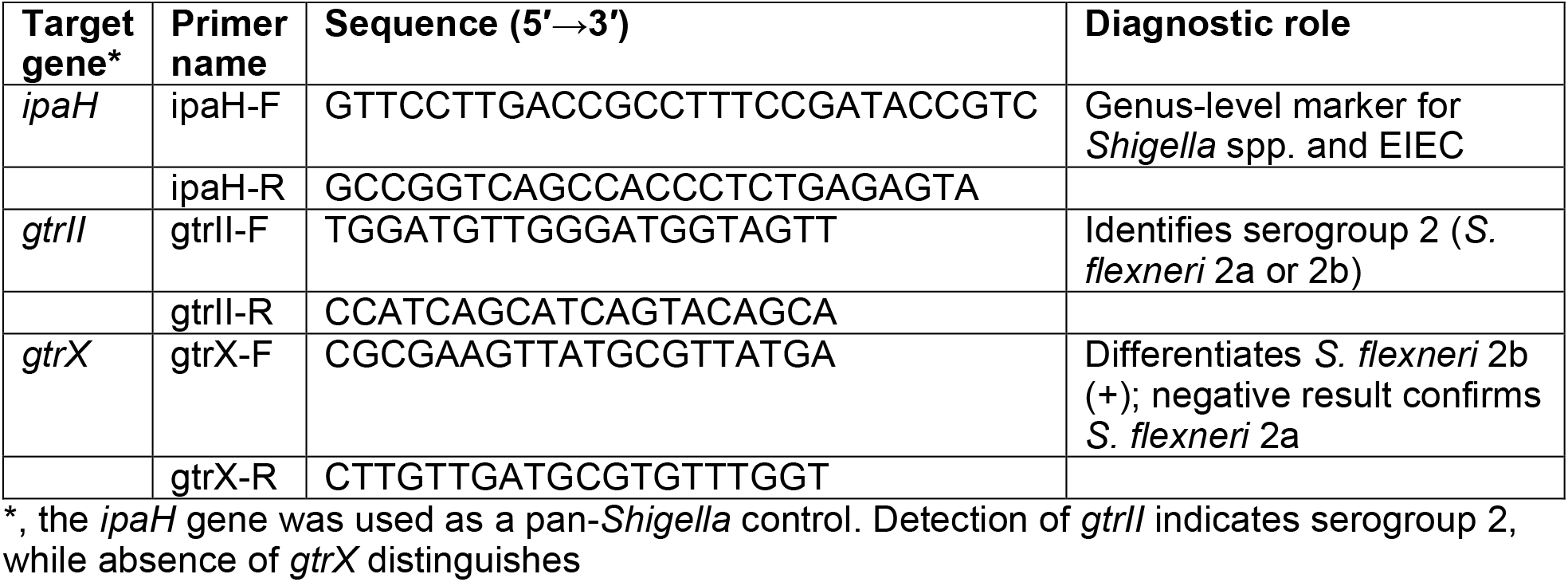
Primer sequences used for qPCR detection of *Shigella flexneri* 2a^19^.

**Supplementary Table 2.** Demographic and clinical characteristics of samples.

| Sample characteristic | Patients (n = 385) | Controls (n = 203) |
| --- | --- | --- |
| Age (mean [SD]), years | 3.5 (± 1.0) | 3.5 (± 1.0) |
| Male (%) | 193 (51.1) | 103 (50.7) |
| Female (%) | 192 (49.9) | 100 (49.3) |
| *Shigella*-positive by standard bacteriological methods* (%) | 385(100) |  |

\*, standard bacteriological methods, including culture and serotyping; SD, standard deviation.

**Supplementary Figure 1.**
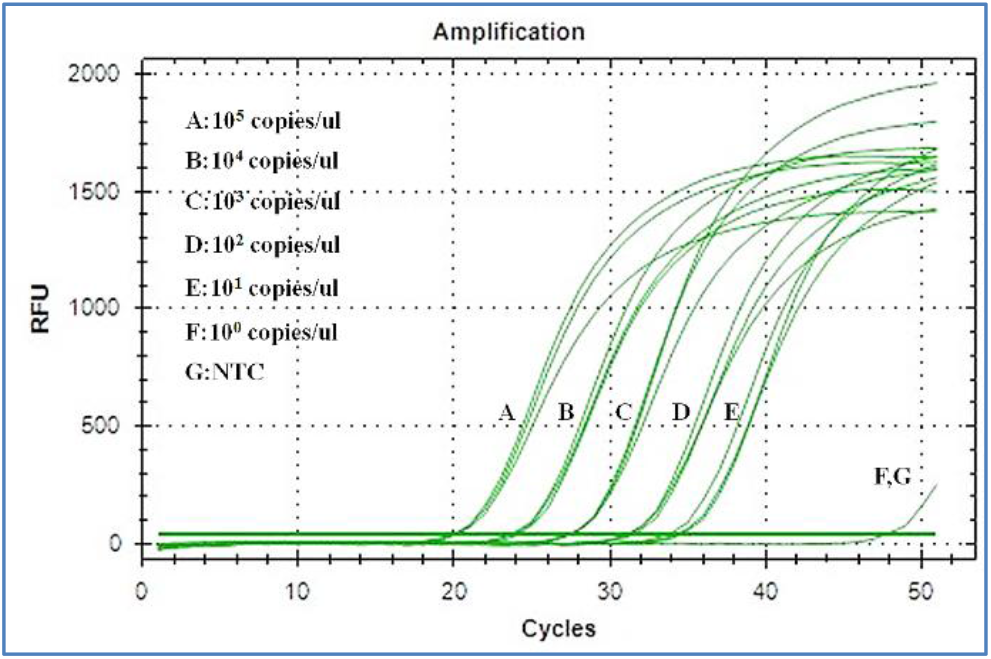
PCR Amplification Curves for Different Copy Numbers of DNA of *S. flexneri*. The concentrations are labeled as follows: A: 10^5^ copies/µl, B: 10^4^ copies/µl, C: 10^3^ copies/µl, D: 10^2^ copies/µl, E: 10^1^ copies/µl, F: 10^0^ copies/µl, and G: NTC (No Template Control). The y-axis represents the relative fluorescence units (RFU), and the x-axis represents the number of PCR cycles. The lowest detectable copy number with a cycle threshold (Ct) of 40 is 10 copies/µl.

